# Adherence to Australian diet, physical activity, and alcohol guidelines is associated with lower risk of depression and anxiety: a secondary, pooled analysis of the CALM trial

**DOI:** 10.1101/2025.10.30.25339194

**Authors:** Lara K Radovic, Rebecca Orr, Melissa M Lane, Felice N Jacka, Tabinda Jabeen, Lauren M Young, Dean Saunders, Sophie Mahoney, Megan Turner, Marita Bryan, Wolfgang Marx, Tetyana Rocks, Adrienne O’Neil, Deborah N Ashtree, the CALM Investigator team

## Abstract

While healthy lifestyle behaviours such as eating a high-quality diet are strongly linked to better mental health, our understanding of how adherence to specific national recommendations in at-risk populations can affect mental health outcomes is limited. This study investigates the relationship between adherence to the Australian diet, physical activity, alcohol use, and smoking guidelines and mental health outcomes in 182 adults experiencing psychological distress. By pooling data from both arms of the CALM trial, a non-inferiority study comparing lifestyle-based therapy to psychotherapy, we examined how adherence to national lifestyle guidelines and specific dietary components over eight weeks was associated with depression and anxiety outcomes. Higher diet quality (RR: 0.93; 0.91), greater physical activity (RR: 0.51; 0.63) and limiting alcohol consumption (RR: 0.59; 0.63) according to guidelines was linked to reduced risk of depression (PHQ-9) and anxiety (GAD-7) respectively, while increased intake of ultra-processed foods was associated with higher risk (RR:1.03; 1.23). Notably, the impact of intake of certain nutrients, such as fibre and omega-6 fatty acids, differed between depression and anxiety outcomes. These findings highlight the importance of integrating lifestyle modification into mental health prevention and treatment strategies and suggest that prescribing dietary behaviours based on national guidelines may offer practical mental health benefits.

## Introduction

Clusters of healthful lifestyle behaviours have consistently been linked with a lower risk of ill mental health (Bourke et al., 2025; Parvin et al., 2025; Tang et al., 2024). These healthy behaviours, such as consuming a good quality diet and adequate high-quality sleep, can have independent and cumulative effects on mental health, where individuals exhibiting the healthiest range of lifestyle habits tend to report fewer ill-health symptoms than all other groups (Bourke et al., 2025). Dietary behaviours are especially closely linked to mental health outcomes (Aucoin et al., 2021; Eliby et al., 2023; Lassale et al., 2019; Meegan et al., 2017; Sadeghi et al., 2021), where a healthier diet has consistently been associated with a reduced risk of depressive symptoms (Aucoin et al., 2024; Guzek et al., 2021; Lassale et al., 2019), while findings for anxiety are less consistent (Aucoin et al., 2021; Basso et al., 2024; Eliby et al., 2023). Other lifestyle behaviours, such as limited alcohol consumption, regular physical activity, and not smoking, are similarly linked to better mental health outcomes (Firth et al., 2020; Kim et al., 2021; Schuch et al., 2018; Schuch et al., 2019; Schuckit, 1996).

Much of the research to date has focused on traditional dietary patterns, particularly the Mediterranean diet, which is strongly linked to lower incidence of depressive symptoms (Altun et al., 2019; Flor-Alemany et al., 2022; Sadeghi et al., 2021). Yet, relatively little attention has been paid to national guidelines, such as the Australian Dietary Guidelines (ADG), which, along with those for alcohol, smoking, and physical activity, are the foundation of nutrition policy and public health messaging in Australia (Australian Government Department of Health, Disability and Ageing, 2013). If considered in the population mental health context, enhancing adherence to ADG has the potential to affect real population-level changes (Hendrie et al., 2022). Where studies have examined guideline adherence, greater individual compliance with national recommendations was associated with better mental health outcomes, both in Australia (Huddy et al., 2016; Lutz et al., 2017; Opie et al., 2020) and internationally (Adjibade et al., 2018; Akbaraly et al., 2013). Still, a clearer understanding of which specific components of these guidelines are most closely associated with mental health benefit is needed, especially in the context of populations with elevated distress. While some studies point to the protective effects of fruit, vegetable, wholegrain, and fish intake, and the harmful impact of some ultra-processed foods (UPFs), such as packaged snacks, sugar-sweetened beverages, and processed meats (Eliby et al., 2023; Lassale et al., 2019), few have tested the impact of these components within national guidelines frameworks (Huddy et al., 2016; Meegan et al., 2017). This knowledge is crucial for refining recommendations and targeting policy interventions that could deliver meaningful mental health improvements at the population level.

Importantly, the ADG recommendations largely align with the dietary risks used by the Global Burden of Disease (GBD) study, which quantifies physical health impacts of modifiable behaviours worldwide (GBD 2019 Diseases and Injuries Collaborators, 2020; GBD 2017 Diet Collaborators, 2019), but these have yet to be applied to mental health outcomes. Recording dietary patterns within the context of national guidelines that align with these GBD risks therefore supports international comparisons, and helps integrate lifestyle risk factors into mental health burden models (GBD Lifestyle and Mental Disorders Taskforce, 2024).

To address these gaps, the current study pools data across both arms of the Curbing Anxiety and depression using Lifestyle Medicine (CALM) trial (O’Neil et al., 2024) — an eight-week, two-arm, parallel-group, non-inferiority trial comparing lifestyle-based therapy (focused on diet, physical activity, and other behaviours) with psychotherapy in adults experiencing elevated distress. This gives us the unique opportunity to examine associations between adherence to the Australian Dietary, Alcohol, Smoking, and Physical Activity Guidelines, specific dietary components, and risk of depression in Australian adults, using validated measures such as the Dietary Guideline Index (DGI; Thorpe, et al. 2016). Our investigation is grounded within the context of the Global Burden of Disease Lifestyle and Mental Disorder (GLAD) project, which was established to facilitate research into the lifestyle factors contributing to depression and anxiety (Ashtree et al., 2025). GLAD provides a critical platform to investigate how diet and lifestyle contributes to mental disorders within a framework aligned with GBD priorities, enabling integration of mental health epidemiology with lifestyle risk modeling. Through this analysis, we can clarify how lifestyle behaviours, individually and collectively, relate to mental health outcomes in an at-risk population.

We expect that: a) greater adherence to Australian lifestyle guidelines will be associated with lower risk of depression and anxiety; b) certain dietary components may show a stronger association with one mental health outcome compared to the other; and c) dietary factors will show independent and cumulative effects (assessed with the DGI). Our findings have the potential to inform both clinical practice and national nutrition and health policy, supporting the integration of dietary and other lifestyle strategies into mental health promotion and treatment. By contributing to the GLAD project and employing a GBD framework, this research contributes to a global, coordinated effort to quantify and mitigate the burden of common mental disorders through targeted lifestyle modifications.

## Methods

### Sample

This study used data from the CALM trial, the details of which have been published elsewhere (O’Neil et al., 2024; Young et al., 2022). Briefly, the CALM trial was a two-arm, parallel-group, non-inferiority trial to determine whether an experimental lifestyle-based therapy was non-inferior to an active control condition (psychotherapy) regarding mental health outcomes and costs when delivered via online videoconferencing.

One-hundred and eighty-two participants were recruited from the Barwon and surrounding regions in Victoria, Australia during the COVID-19 lockdowns between May 2021 and April 2022. Participants were included if they were: aged 18 years or over; able to provide written informed consent; proficient in English; able to attend six 90-minute sessions over an eight-week period; digitally literate; and deemed to be a likely depression case, based on a cut-off of ≥8 on the Distress Questionnaire-5 (Batterham et al., 2017). Participants were excluded if they had: a clinically unstable medical disorder; a formally diagnosed eating disorder; severe dietary allergies; socio-cultural, religious, or medical reasons preventing participation; enrolled in another trial; or a current or planned pregnancy or breastfeeding.

The lifestyle program was co-developed and co-delivered by Accredited Practising Dietitians and Exercise Physiologists. The content of these sessions focused on setting relevant goals to achieve positive lifestyle change, mostly relating to nutrition and physical activity, however participants could also nominate other lifestyle targets (e.g. alcohol, smoking, substance use, sleep hygiene). The nutritional counselling was derived from the Mod*i*Med Diet and did not target weight change.

The psychotherapy program used a Cognitive Behavioural Therapy (CBT) approach, co-facilitated by two psychologists. Content included skills to promote self-awareness, identifying and managing unhelpful behaviours and thoughts, and mindfulness practices.

### Exposures

Dietary intake was measured using the Dietary Questionnaire for Epidemiological Studies version 3.2 (DQES v3.2) at baseline and eight-week follow-up. The DQES v3.2 is a self-reported food frequency questionnaire developed by the Cancer Council Victoria (CCV, 2018) and has been validated against weighed food records (Hebden et al., 2013). Participants self-reported dietary consumption of 72 food items.

Our primary exposure of interest was the revised DGI (Thorpe, et al. 2016), a measure of overall diet quality and how well participants adhered to the Australian Dietary Guidelines (ADG; Australian Government Department of Health, Disability and Ageing, 2013). Higher scores indicated greater adherence and a total possible score of 120. The key features of the ADG include: 1) achieving and maintaining a healthy weight, being physically active and choosing amounts of nutritious food and drinks to meet energy needs; 2) enjoying a wide variety of nutritious foods from five groups (vegetables, including different types and colours, and legumes/beans; fruit; grains, mostly wholegrain and/or high fibre varieties; lean meats and alternatives, including poultry, fish, eggs, tofu, nuts and seeds, and legumes/beans; and milk, yoghurt, cheese and their alternatives); 3) drink plenty of water; and 4) limit intake of foods containing saturated fat, added salt, added sugars and alcohol.

We were additionally interested in identifying whether specific components of dietary intake were associated with greater or reduced risk of depression and anxiety over eight weeks. The dietary components were defined according to dietary risks used by the GBD study (GBD 2019 Risk Factors Collaborators, 2020; GBD 2021 Risk Factors Collaborators, 2024) and included the following variables: fruit (g/day), vegetables (excluding starchy vegetables) (g/day), legumes (g/day), wholegrains (g/day), nuts and seeds (g/day), dairy-based milk (g/day), red meat (g/day), processed meat (g/day), sugar-sweetened beverages (g/day), fibre (g/day), calcium (mg/day), polyunsaturated fat (% energy/day), omega-3 (g/day), omega-6 (% energy/day) and sodium (g/day). We additionally included UPF intake (g/day), based on the Nova food classification system (Monteiro et al., 2019) due to increasing evidence linking UPF intake to a range of health outcomes (Lane et al., 2024).

In addition to these dietary measures, we include adherence to Australian physical activity, alcohol consumption, and smoking guidelines in this analysis (Australian Government Department of Health, Disability and Ageing, 2020; 2021). We determined whether physical activity habits met guidelines through information provided by participants through the Simple Physical Activity Questionnaire (Rosenbaum et al., 2020) at eight-week follow-up, while information on alcohol consumption was obtained through the DQES v3.2, and current/former smokers were identified through information provided in the WHO-ASSIST (Humeniuk et al., 2010) questionnaire at this same time point.

### Outcomes

Our primary outcome of interest was ‘likely depression’ diagnosis, measured at the eight-week follow-up via the nine-item Patient Health Questionnaire (PHQ-9). A cut-off score of 10 was used to identify likely depression (Kroenke, Spitzer, & Williams, 2001). We additionally included ‘likely anxiety’ diagnosis as a secondary outcome. Anxiety was measured at eight weeks using the generalised anxiety disorder seven item scale (GAD-7). We used a cut-off score of 10 on the GAD-7 to identify anxiety diagnosis (Spitzer et al., 2006).

### Covariates

At the baseline intake interviews, CALM participants reported demographic information, including sex, age, and postcodes. Postcodes were used to obtain area-level socioeconomic status, the Socioeconomic Index for Areas Index of Relative Socioeconomic Advantage and Disadvantage (SEIFA-IRSAD). SEIFA-IRSAD scores are calculated by the Australian Bureau of Statistic to measure relative socioeconomic advantage of given geographical areas at the postcode-level (Australian Bureau of Statistics, 2023). To remove the potential confounding effect of energy intake, we adjusted for energy using Willett’s residual method (Willett, Howe, & Kushi, 1997).

### Statistical Analyses

The CALM trial found no difference between the two treatment arms with respect to depression and anxiety outcomes, or with fruit, vegetable, grains, dairy, fibre, omega-3, omega-6 or polyunsaturated fats (O’Neil et al., 2024). Therefore, to maximise statistical power, our initial model combined both treatment arms to longitudinally assess the association of each dietary variable with risk of depression and anxiety. A generalised estimating equation Poisson regression model with robust standard errors was used to account for the group-based nature of the intervention and the time-varying aspect of the diet (due to dietary goals being part of the lifestyle treatment arm). Two models were fitted for each dietary exposure and outcome pairing: 1) adjusted for energy intake only; and 2) adjusting for age, sex, socioeconomic status, and total energy intake. We conducted an additional subgroup model to determine whether results differed by treatment arm (see supplementary material for full results). Intention-to-treat (ITT) analyses were conducted, whereby missing data were imputed using multiple imputation chained equations. Magnitudes of effect were presented as risk ratios with accompanying 95% Confidence Intervals (95% CIs). We used the Simes method of p-value adjustment to account for multiple testing (Simes, 1986).

### Ethics

The CALM trial was approved by the Barwon Health (20/199) and Deakin University (2021-166) Human Research Ethics Committees (HREC). The trial was conducted in accordance with principles of the Declaration of Helsinki, Australian National Health and Medical Research Council National Statement on Ethical Conduct in Human Research (2007) and the Note for Guidance on Good Clinical Practice (CPMP/ICH-135/95). All participants provided written informed consent. The trial was registered with the Australian New Zealand Clinical Trials Registry (Registry number: ACTRN12621000387820).

This manuscript has been prepared in accordance with the requirements of the GLAD Taskforce, as part of a global collaborative project to inform the GBD, Injuries, and Risk Factors Study for which the protocol is available (Ashtree et al., 2025).

## Results

Table 1 below summarises the demographics and lifestyle behaviours of the overall sample and by arm. No difference was found between treatment arms for depression and anxiety outcomes, or intake of fruits, vegetables, grains, dairy, fibre, omega-3s, omega-6s or polyunsaturated fats.

**Table 1:**
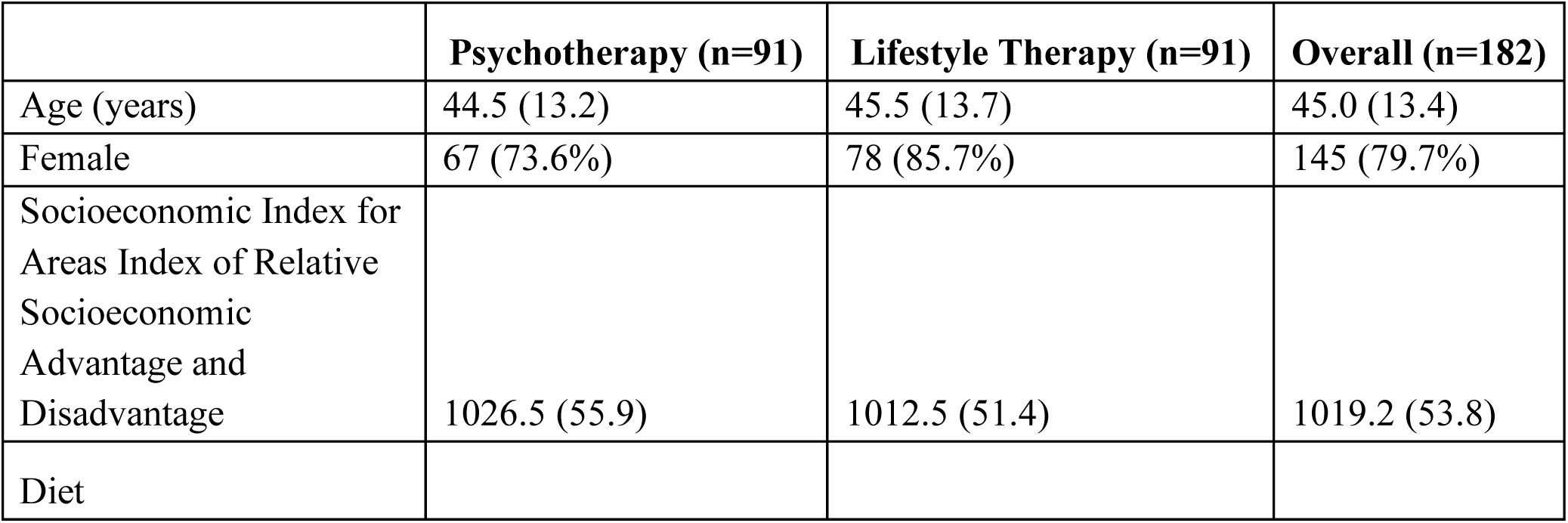
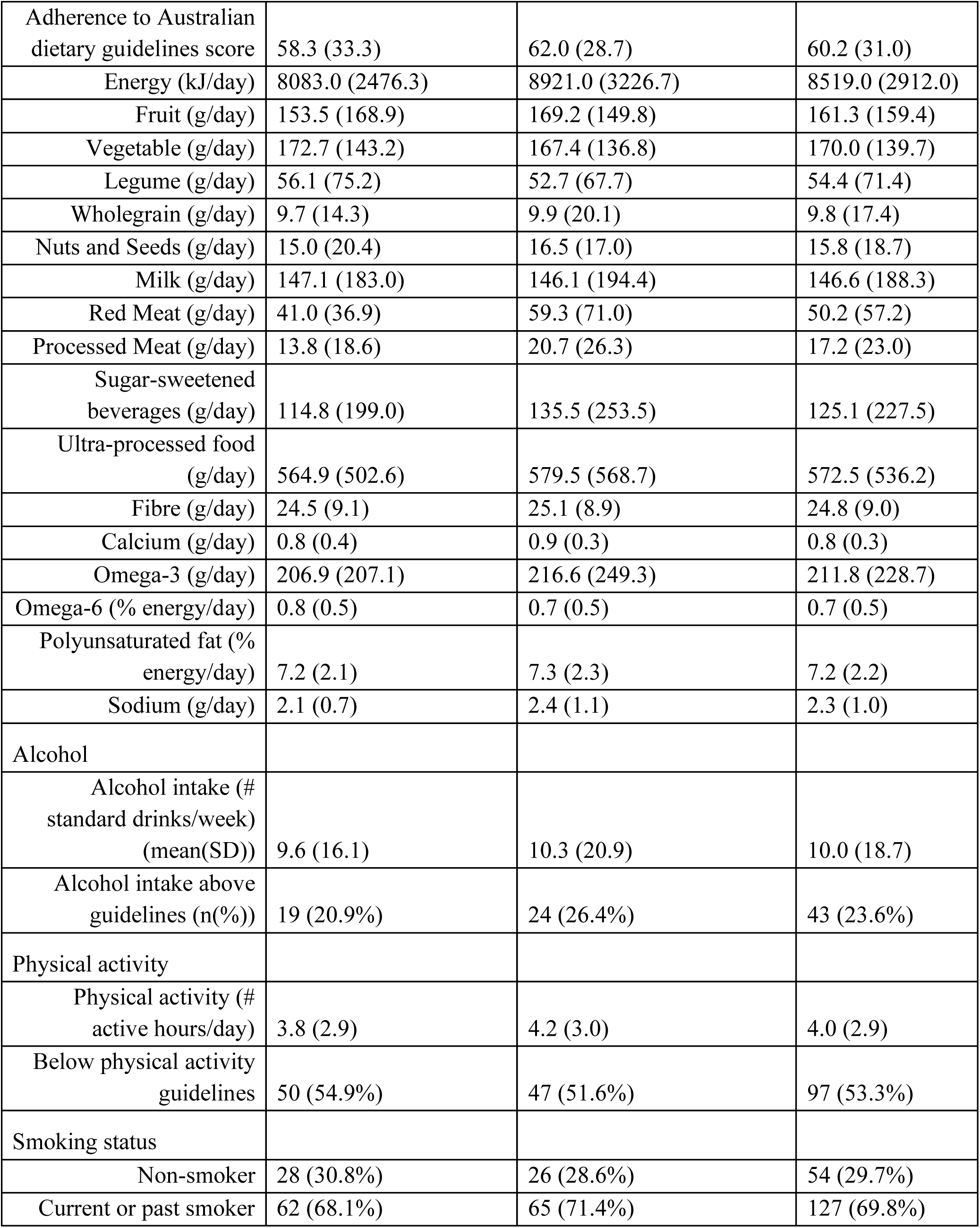
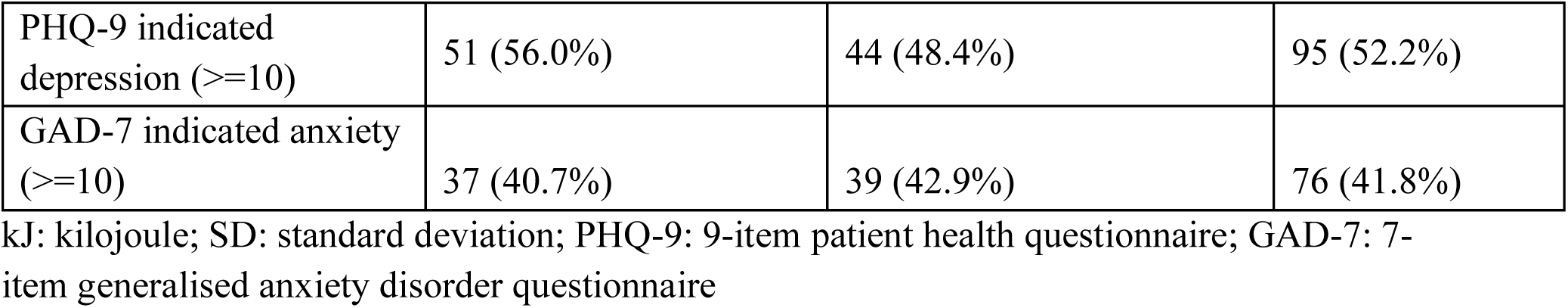
Demographics and Lifestyle Measures of the CALM sample.

|  | **Psychotherapy (n=91)** | **Lifestyle Therapy (n=91)** | **Overall (n=182)** |
| --- | --- | --- | --- |
| Age (years) | 44.5 (13.2) | 45.5 (13.7) | 45.0 (13.4) |
| Female | 67 (73.6%) | 78 (85.7%) | 145 (79.7%) |
| Socioeconomic Index for Areas Index of Relative Socioeconomic Advantage and Disadvantage | 1026.5 (55.9) | 1012.5 (51.4) | 1019.2 (53.8) |
| Diet |  |  |  |
| Adherence to Australian dietary guidelines score | 58.3 (33.3) | 62.0 (28.7) | 60.2 (31.0) |
| Energy (kJ/day) | 8083.0 (2476.3) | 8921.0 (3226.7) | 8519.0 (2912.0) |
| Fruit (g/day) | 153.5 (168.9) | 169.2 (149.8) | 161.3 (159.4) |
| Vegetable (g/day) | 172.7 (143.2) | 167.4 (136.8) | 170.0 (139.7) |
| Legume (g/day) | 56.1 (75.2) | 52.7 (67.7) | 54.4 (71.4) |
| Wholegrain (g/day) | 9.7 (14.3) | 9.9 (20.1) | 9.8 (17.4) |
| Nuts and Seeds (g/day) | 15.0 (20.4) | 16.5 (17.0) | 15.8 (18.7) |
| Milk (g/day) | 147.1 (183.0) | 146.1 (194.4) | 146.6 (188.3) |
| Red Meat (g/day) | 41.0 (36.9) | 59.3 (71.0) | 50.2 (57.2) |
| Processed Meat (g/day) | 13.8 (18.6) | 20.7 (26.3) | 17.2 (23.0) |
| Sugar-sweetened beverages (g/day) | 114.8 (199.0) | 135.5 (253.5) | 125.1 (227.5) |
| Ultra-processed food (g/day) | 564.9 (502.6) | 579.5 (568.7) | 572.5 (536.2) |
| Fibre (g/day) | 24.5 (9.1) | 25.1 (8.9) | 24.8 (9.0) |
| Calcium (g/day) | 0.8 (0.4) | 0.9 (0.3) | 0.8 (0.3) |
| Omega-3 (g/day) | 206.9 (207.1) | 216.6 (249.3) | 211.8 (228.7) |
| Omega-6 (% energy/day) | 0.8 (0.5) | 0.7 (0.5) | 0.7 (0.5) |
| Polyunsaturated fat (% energy/day) | 7.2 (2.1) | 7.3 (2.3) | 7.2 (2.2) |
| Sodium (g/day) | 2.1 (0.7) | 2.4 (1.1) | 2.3 (1.0) |
| Alcohol |  |  |  |
| Alcohol intake (# standard drinks/week) (mean(SD)) | 9.6 (16.1) | 10.3 (20.9) | 10.0 (18.7) |
| Alcohol intake above guidelines (n(%)) | 19 (20.9%) | 24 (26.4%) | 43 (23.6%) |
| Physical activity |  |  |  |
| Physical activity (# active hours/day) | 3.8 (2.9) | 4.2 (3.0) | 4.0 (2.9) |
| Below physical activity guidelines | 50 (54.9%) | 47 (51.6%) | 97 (53.3%) |
| Smoking status |  |  |  |
| Non-smoker | 28 (30.8%) | 26 (28.6%) | 54 (29.7%) |
| Current or past smoker | 62 (68.1%) | 65 (71.4%) | 127 (69.8%) |
| PHQ-9 indicated depression ($\geq 10$) | 51 (56.0%) | 44 (48.4%) | 95 (52.2%) |
| GAD-7 indicated anxiety ($\geq 10$) | 37 (40.7%) | 39 (42.9%) | 76 (41.8%) |

kJ: kilojoule; SD: standard deviation; PHQ-9: 9-item patient health questionnaire; GAD-7: 7-item generalised anxiety disorder questionnaire

### Associations between diet, physical activity, alcohol intake, and depression

After adjusting for energy intake, sex, age, and socioeconomic status in the ITT analyses, for every 10-unit increase in diet quality index score, risk of depression decreased 0.93-fold (95%CI=0.89-0.97; Figure 1). Adherence to the Australian physical activity guidelines was also associated with a lower risk of depression (RR:0.51, 95%CI=0.38-0.67), along with adherence to alcohol guidelines (RR: 0.59, 95%CI=0.41-0.84). There was no difference in depression risk for non-smokers compared to current or former smokers.

**Figure 1:**
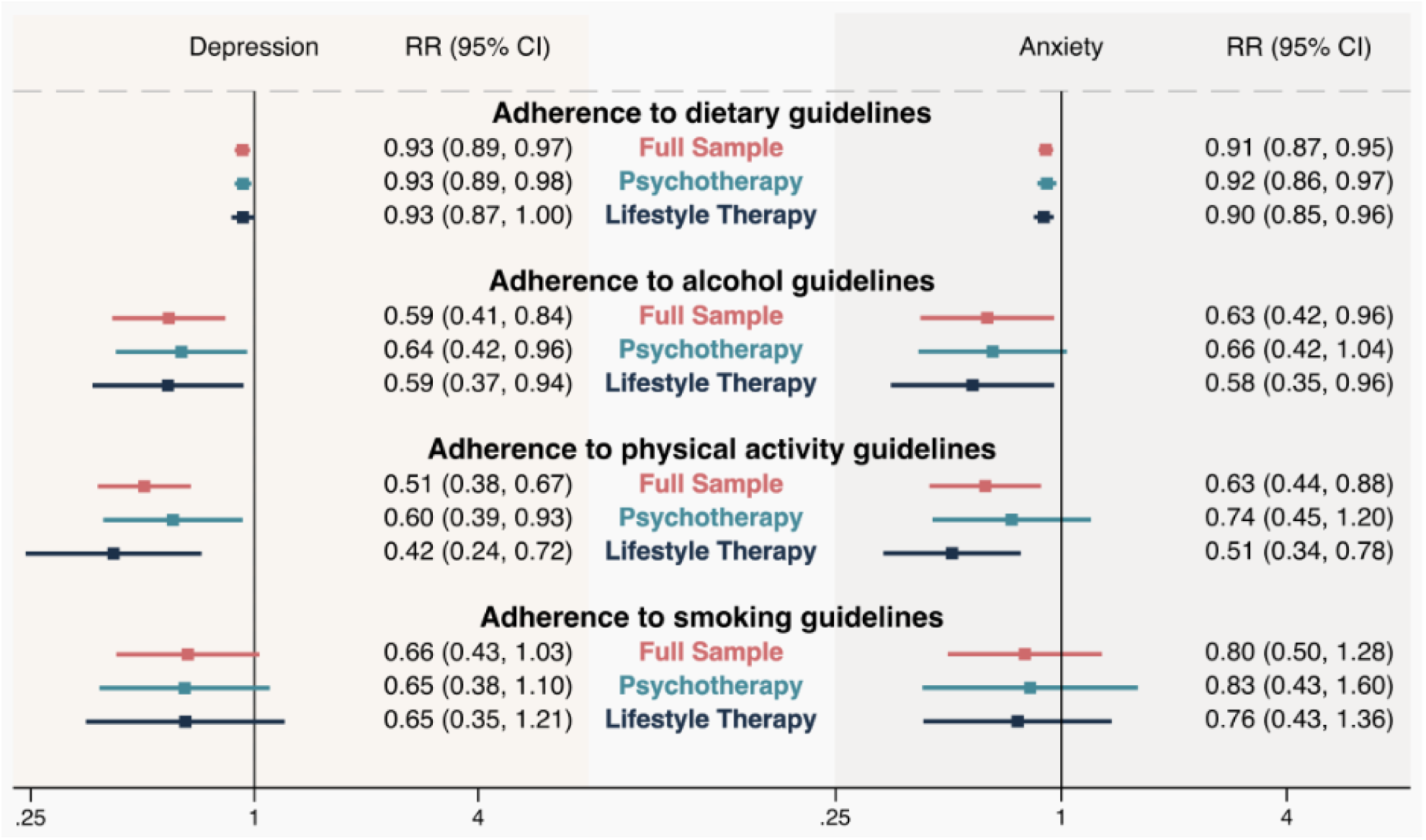
Association of adherence to Australian guidelines with anxiety and depression in the CALM trial after adjusting for energy intake, sex, age and SES.

When looking at specific lifestyle components, after adjusting for energy intake, sex, age, and socioeconomic status, higher vegetable intake (RR: 0.92 per 75g increase, 95% CI=0.87-0.98), nuts and seeds intake (RR: 0.80 per 30g increase, 95%CI=0.65-0.99), fibre intake (RR=0.68 per 30g increase, 95%CI=0.50-0.94), and physical activity (RR: 0.93 per 1hr/week increase, 95%CI=0.88-0.98) were associated with lower risk of depression (Table 2).

**Table 2:**
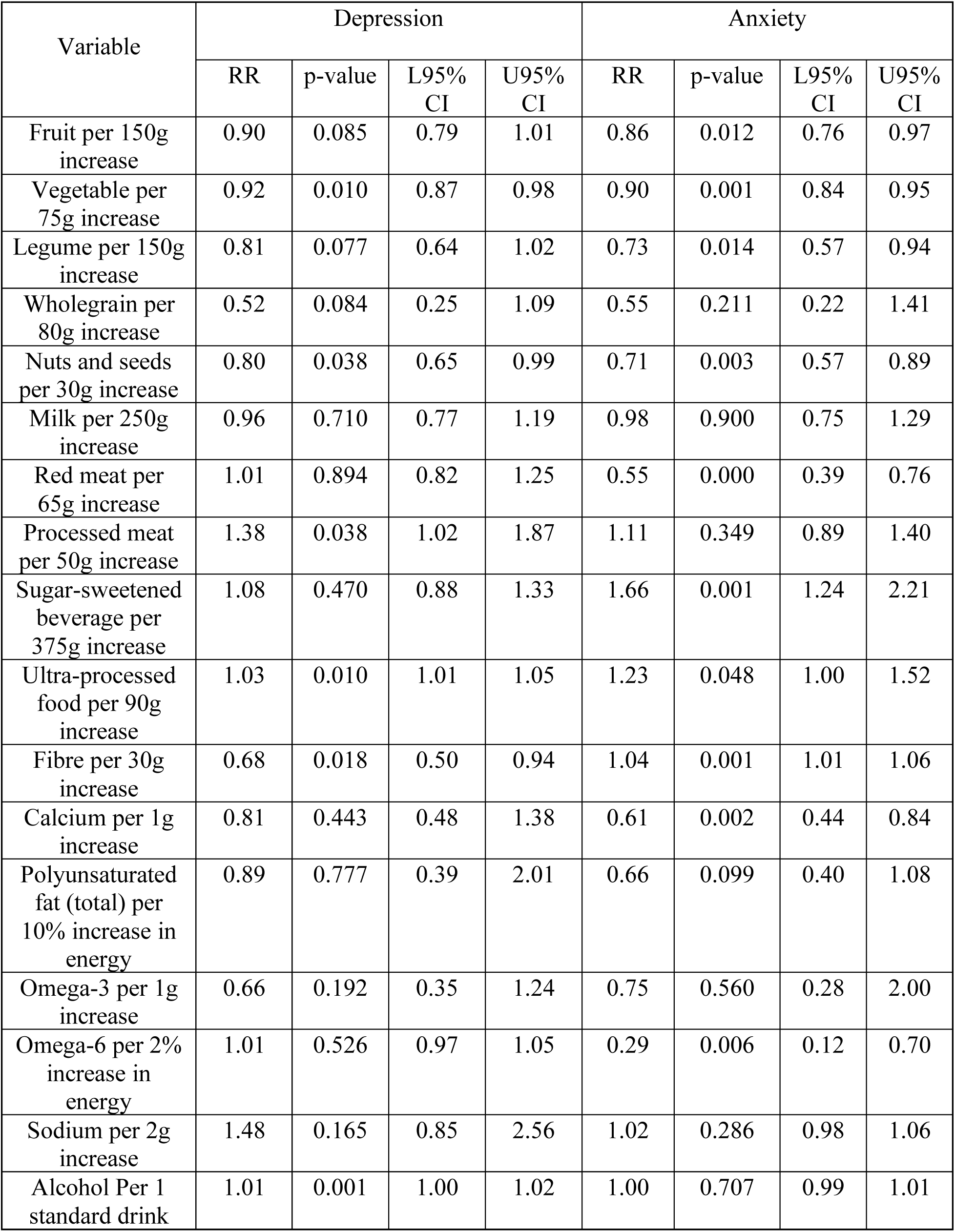
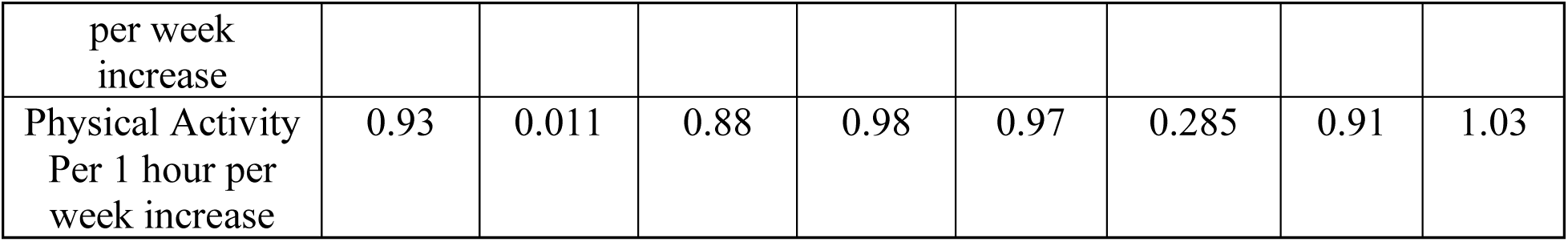
Association of lifestyle variables with risk of depression and anxiety over 8 weeks after adjusting for energy intake, sex, age, and socioeconomic status.

Conversely, the overall UPF intake (RR: 1.03 per 90g increase, 95%CI=1.01-1.05), processed meat intake (RR: 1.38 per 50g increase, 95%CI=1.02-1.87), and alcohol intake (RR: 1.01 per 1 standard drink increase, 95%CI=1.00-1.02) were associated with higher risk of depression.

### Associations between diet, physical activity, alcohol intake, and anxiety

After adjusting for energy intake, sex, age, and socioeconomic status in the ITT analyses, for every 10 unit increase in diet quality index score, risk of anxiety decreased 0.91-fold (95%CI=0.87-0.95; Figure 1). Adherence to the Australian physical activity guidelines was also associated with a lower risk of anxiety (RR:0.63, 95%CI=0.44-0.88), along with adherence to alcohol guidelines (RR: 0.63, 95%CI=0.42-0.96). There was no difference in anxiety risk for non-smokers compared to current or former smokers.

When looking at specific lifestyle components, after adjusting for age, sex, socioeconomic status, and energy intake, higher intake of UPFs (RR: 1.23 per 90g increase, 95%CI=1.00-1.52), sugar-sweetened beverages (RR: 1.66 per 375g increase, 95%CI=1.24-2.21) and, surprisingly, fibre (RR: 1.04 per 30g increase, 95%CI=1.01-1.06) were associated with increased risk of anxiety (Table 2). Higher intake of fruit (RR: 0.86 per 150g increase, 95%CI=0.76-0.97), vegetables (RR= 0.90 per 75g increase, 95%CI=0.84-0.95), legumes (RR: 0.73 per 150g increase, 95%CI=0.57-0.94), nuts and seeds (RR: 0.71 per 30g increase, 95%CI=0.57-0.89), red meat (RR= 0.55 per 65g increase, 95%CI=0.39-0.76), calcium (RR= 0.61 per 1g increase, 95%CI=0.44-0.84), and omega-6s (RR= 0.29 per 2% of energy increase, 95%CI=0.12-0.70) was associated with lower risk of anxiety.

## Discussion

This study contributes new evidence about the role of lifestyle behaviours, including diet, physical activity, alcohol consumption, and smoking, in the management of depression and anxiety in a sample with elevated psychological distress. Our findings align with prior research showing that higher adherence to national dietary and physical activity guidelines, as well as specific dietary components, is associated with reduced risk of depressive and anxiety symptoms (Aucoin et al., 2021; Eliby et al., 2023; Lassale et al., 2019; Schuch et al., 2018). Here we demonstrated that these associations are present within an ‘at risk’ population (i.e., with elevated psychological distress) undergoing active intervention, thus supporting the relevance of lifestyle modification in treatment settings.

Consistent with earlier cohort and intervention studies, our data show that higher diet quality measured by the DGI was associated with a lower risk of both depression and anxiety. This echoes reports of adherence to healthy dietary patterns protecting against or improving depression in various populations (Eliby et al., 2023; Lassale et al., 2019). Similarly, our results extend work exploring international and national lifestyle guidelines (Adjibade et al., 2018; Huddy et al., 2016; Opie et al., 2020), finding that adherence predicts better mental health outcomes. While past research has focused largely on Mediterranean or similar dietary patterns (Altun et al., 2019; Bayes et al., 2022), our findings further support the mental health benefits of following relevant national guidelines.

At the dietary component level, our study found that greater intakes of vegetables, nuts and seeds, and fibre were associated with reduced risk of depression, consistent with earlier work highlighting the value of these and other whole food consumption for mental health (Guzek et al., 2021; Meegan et al., 2017). The association between higher fruit, vegetable, legume, and nut intake and lower risk of anxiety also aligns with emerging evidence (Basso et al., 2024; Eliby et al., 2023). The observed protective association of red meat, calcium, and omega-6 intake on anxiety risk is less commonly reported and warrants further study. Conversely, higher consumption of UPFs and alcohol were associated with greater risk of depression and anxiety, in line with existing findings (Kim et al., 2021; Lane et al., 2024).

Differences between the dietary components found to be linked to depression and anxiety respectively suggest that certain nutrients may have distinct influences. Higher fibre intake, for example, was associated with a lower risk of depression but higher risk of anxiety. While the beneficial effects of fibre on gut microbiota and systemic inflammation may protect against depression (Dalile et al., 2019; Marx et al., 2021), its relationship with anxiety could be more complex, potentially reflecting differences in gut-brain axis signaling or sensitivity to fibres that influence anxiety symptoms. Still, studies to date all show an inverse association between fibre consumption and anxiety symptoms (Burokas et al., 2017; Saghafian et al., 2021). A potential explanation for our contradictory finding is that individuals with elevated anxiety, particularly health-related worry, may become more health conscious and alter their diets accordingly, similarly to studies conducted during COVID-19 that report a link between higher anxiety and increased preventative health behaviours (Stefan et al., 2021). Additionally, intakes of fruit, legumes, red meat, calcium, and omega-6 fatty acids were associated with reduced anxiety risk but showed no association with depression. These findings could mean that anxiety symptoms are particularly sensitive to certain micronutrients, potentially through nutrient-driven modulation of neurotransmitters. For example, calcium plays a crucial role in neuronal excitability and synaptic function (Marambaud et al., 2009), while omega-6 fatty acids are precursors to signaling molecules, though a lower omega-6:omega-3 fatty acid ratio is typically considered favourable for mood (Pinto et al., 2025). However, linoleic acid (the primary omega-6 fatty acid) is not always converted to its inflammatory derivative (Su et al., 2017), and higher intake of omega-6s could simply be an indicator of an overall healthy diet (high in nuts, seeds, and plant oils rather than saturated fats). The lack of association between the above components and depression found in our study may reflect differing underlying neurobiological mechanisms between the disorders, suggesting a potential need for targeted exploration to identify specific differences in nutritional strategies for anxiety and depression. Further research is warranted to confirm whether modifying intake of these nutrients could differentially benefit anxiety and depression and how this approach could be relevant for those with these comorbid conditions.

Physical activity was also associated with lower risk of depression and anxiety, consistent with prospective studies and meta-analyses demonstrating preventive and therapeutic effects of exercise (Firth et al., 2020; Kim et al., 2021; Schuch et al., 2018). Adherence to alcohol guidelines was similarly protective, supporting prior research linking excessive alcohol use to greater depression and anxiety (Kim et al., 2021; Schuckit, 1996). However, unlike many large-scale epidemiological studies reporting a significant association between smoking and poor mental health outcomes (Firth et al., 2020; Kim et al., 2021; Pasco et al., 2008), our study did not provide evidence for a difference in depression or anxiety symptoms between non-smokers and current or former smokers. Given that our sample had a substantial number of current/former smokers, this discrepancy is unlikely due to sample size. Instead, it may reflect a limitation in how the measure is defined (without including detailed smoking history (e.g., duration, intensity)).

### Mechanisms and Interpretations

Several biological and psychosocial mechanisms may underlie the observed associations between lifestyle behaviors and mental health outcomes. Diets rich in plant-based foods, fibre, and healthy fats may reduce risk of depression and anxiety through anti-inflammatory and neuroprotective effects, modulation of gut microbiota, and enhancements in neuroplasticity (Marx et al., 2021). Conversely, the detrimental impact of UPFs, including processed meats and sugar-sweetened beverages, may be mediated by increased systemic inflammation and dysregulation of neurotransmitter systems (Lane et al, 2024; Lutz et al., 2025). The protective association of red meat and calcium intake on anxiety found in this sample is less established in the literature and may reflect adequate micronutrient intake, particularly iron, zinc, and B vitamins, which improve neurocognitive health (Parletta et al., 2013; Tardy et al., 2020).

Physical activity can also benefit mental health through reductions in inflammation, hypothalamic-pituitary-adrenal axis (stress) regulation, and increased neurogenesis (Hossain et al., 2024). The relationship between alcohol and mental health is complex, but excessive intake may exacerbate mood symptoms through neurochemical and psychosocial mechanisms including neurotransmitter imbalance and social stressors (Kim et al., 2021; Schuckit, 1996; Valenzuela et al., 1997).

### Limitations

Several limitations must be considered when interpreting these results. Firstly, though based on a randomised trial, this secondary analysis is observational in nature and therefore precludes causal inference. Given this was a sample at risk of mental disorders (at least elevated distress at enrolment), mental health may have influenced lifestyle behaviours. Secondly, dietary and lifestyle behaviors were self-reported, which may introduce recall or social desirability bias. Thirdly, the sample was recruited during the COVID-19 pandemic and from a specific region in Australia, which limits generalisability. There was also a high volume of missing data, making imputation and use of an ITT model necessary, leading to potential bias in our findings.

### Future Directions

Future research should replicate these findings in larger, more diverse cohorts with longer follow-up to better understand the nature of these associations, as well as any long-term effects of lifestyle behaviours on mental health outcomes over time. Studies incorporating more detailed measures of smoking status, alcohol consumption, and physical activity within their analyses, as we did here with dietary components relevant to the GBD study, would improve confidence in these associations. Intervention trials targeting specific dietary components and physical activity, along with substance use reduction, could clarify causality and optimise treatment strategies.

Integration of lifestyle data into global mental health burden models, as facilitated by initiatives like GLAD, remains a priority for informing policy.

### Implications

These findings emphasise the potential value of integrating lifestyle modification, particularly improvements in diet quality and physical activity, into mental health treatment strategies, especially in at-risk populations. Adherence to national guidelines represents a practical and effective target for clinical and public health interventions that have a real potential to contribute to lowering mental disorder incidence. Given the global burden of depression and anxiety, even modest improvements in population-level lifestyle behaviors could translate into meaningful mental health gains.

## Data Availability

A minimum dataset (including biological specimens) of de-identified individual participant data collected during the trial is available (including data dictionaries, study protocol, statistical analysis plan, analytic code) for any purpose that is approved by the Barwon Health Human Research in Ethics Committee and the CALM Chief Investigator Team.

https://researchdata.edu.au/health/

## Acknowledgements

The Community and Research Network (CARN) and Elizabeth Elms for input into program design and delivery, the participants who volunteered their time for this study and all team members who worked on the CALM trial.

## Notes

**Funding** This work was supported by the National Health and Medical Research Council’s Medical Research Future Fund under the COVID-19 Mental Health Research Australian Government Department of Health grant (GA133346). LKR, RO, and TJ are supported by a Deakin University Postgraduate Research Scholarship. AON and DNA are supported by a National Health and Medical Research Council Emerging Leader 2 Fellowship (#2009295). FNJ is supported by a National Health and Medical Research Council Leader 1 Fellowship (#1194982). MML is supported by Deakin University Postdoctoral Fellowships. TR is supported by the Roberts Family foundation.

### Competing Interest Statement

The authors have declared no competing interest.

### Clinical Trial

ACTRN12621000387820

### Funding Statement

This work was supported by the National Health and Medical Research Council Medical Research Future Fund under the COVID-19 Mental Health Research Australian Government Department of Health grant (GA133346).

### Author Declarations

The CALM trial was approved by the Barwon Health (20/199) and Deakin University (2021-166) Human Research Ethics Committees (HREC). The trial was conducted in accordance with principles of the Declaration of Helsinki, Australian National Health and Medical Research Council National Statement on Ethical Conduct in Human Research (2007) and the Note for Guidance on Good Clinical Practice (CPMP/ICH-135/95). All participants provided written informed consent.This manuscript has been prepared in accordance with the requirements of the GLAD Taskforce, as part of a global collaborative project to inform the GBD, Injuries, and Risk Factors Study for which the protocol is available.

