## Supplemental Tables for "Adherence to Australian diet, physical activity, and alcohol guidelines is associated with lower risk of depression and anxiety: a secondary, pooled analysis of the CALM trial"

### Supplementary Materials

#### Tables 3ab: *Association of adherence to Australian guidelines with risk of depression and anxiety over 8 weeks by arm*

| Depression | Full Sample: Unadjusted | | | | | Full Sample: Adjusted for age, sex and SES | | | | | Psychotherapy: Unadjusted | | | | | Psychotherapy: Adjusted for age, sex, and SES | | | | | Lifestyle Therapy: Unadjusted | | | | | Lifetyle Therapy: Adjusted for age, sex, and SES | | | | |
|---|---|---|---|---|---|---|---|---|---|---|---|---|---|---|---|---|---|---|---|---|---|---|---|---|---|---|---|---|---|---|
| | RR | P>\|t\| | Q | L95% CI | U95% CI | RR | P>\|t\| | Q | L95% CI | U95% CI | RR | P>\|t\| | Q | L95% CI | U95% CI | RR | P>\|t\| | Q | L95% CI | U95% CI | RR | P>\|t\| | Q | L95% CI | U95% CI | RR | P>\|t\| | Q | L95% CI | U95% CI |
| Adherence to Australian dietary guidelines (per 10 unit increase) | 0.93 | 0.001 | 0.003 | 0.89 | 0.97 | 0.93 | 0.002 | 0.008 | 0.89 | 0.97 | 0.93 | 0.001 | 0.003 | 0.89 | 0.97 | 0.93 | 0.008 | 0.019 | 0.89 | 0.98 | 0.93 | 0.042 | 0.055 | 0.87 | 1.00 | 0.93 | 0.054 | 0.072 | 0.87 | 1.00 |
| Adherence to Australian alcohol guidelines (met/below guidelines vs exceeded guidelines) | 0.61 | 0.003 | 0.007 | 0.43 | 0.85 | 0.59 | 0.003 | 0.009 | 0.41 | 0.84 | 0.61 | 0.012 | 0.024 | 0.42 | 0.90 | 0.64 | 0.030 | 0.045 | 0.42 | 0.96 | 0.64 | 0.046 | 0.055 | 0.41 | 0.99 | 0.59 | 0.025 | 0.043 | 0.37 | 0.94 |
| Adherence to Australian physical activity guidelines (met/exceeded guidelines vs below guidelines) | 0.50 | 0.000 | 0.000 | 0.37 | 0.67 | 0.51 | 0.000 | 0.000 | 0.38 | 0.67 | 0.60 | 0.027 | 0.046 | 0.38 | 0.94 | 0.60 | 0.022 | 0.043 | 0.39 | 0.93 | 0.42 | 0.001 | 0.003 | 0.24 | 0.71 | 0.42 | 0.002 | 0.008 | 0.24 | 0.72 |
| Adherence to Australian smoking guidelines (non-smokers vs current/former smokers) | 0.63 | 0.057 | 0.062 | 0.39 | 1.01 | 0.66 | 0.069 | 0.083 | 0.43 | 1.03 | 0.57 | 0.032 | 0.048 | 0.34 | 0.95 | 0.65 | 0.108 | 0.118 | 0.38 | 1.10 | 0.67 | 0.231 | 0.231 | 0.34 | 1.29 | 0.65 | 0.173 | 0.173 | 0.35 | 1.21 |

| Anxiety | Full Sample: Unadjusted | | | | | Full Sample: Adjusted for age, sex and SES | | | | | Psychotherapy: Unadjusted | | | | | Psychotherapy: Adjusted for age, sex, and SES | | | | | Lifestyle Therapy: Unadjusted | | | | | Lifestyle: Adjusted for age, sex, and SES | | | | |
|---|---|---|---|---|---|---|---|---|---|---|---|---|---|---|---|---|---|---|---|---|---|---|---|---|---|---|---|---|---|---|
| | RR | P>\|t\| | Q | L95% CI | U95% CI | RR | P>\|t\| | Q | L95% CI | U95% CI | RR | P>\|t\| | Q | L95% CI | U95% CI | RR | P>\|t\| | Q | L95% CI | U95% CI | RR | P>\|t\| | Q | L95% CI | U95% CI | RR | P>\|t\| | Q | L95% CI | U95% CI |
| Adherence to Australian dietary guidelines (per 10 unit increase) | 0.91 | 0.000 | 0.000 | 0.87 | 0.95 | 0.91 | 0.000 | 0.000 | 0.87 | 0.95 | 0.92 | 0.001 | 0.004 | 0.88 | 0.97 | 0.92 | 0.003 | 0.009 | 0.86 | 0.97 | 0.90 | 0.000 | 0.000 | 0.84 | 0.95 | 0.90 | 0.001 | 0.006 | 0.85 | 0.96 |
| Adherence to Australian alcohol guidelines (met/below guidelines vs exceeded guidelines) | 0.66 | 0.024 | 0.048 | 0.46 | 0.95 | 0.63 | 0.030 | 0.057 | 0.42 | 0.96 | 0.69 | 0.049 | 0.080 | 0.48 | 1.00 | 0.66 | 0.071 | 0.107 | 0.42 | 1.04 | 0.62 | 0.053 | 0.080 | 0.38 | 1.01 | 0.58 | 0.033 | 0.057 | 0.35 | 0.96 |
| Adherence to Australian physical activity guidelines (met/exceeded guidelines vs below guidelines) | 0.63 | 0.009 | 0.022 | 0.45 | 0.89 | 0.63 | 0.008 | 0.019 | 0.44 | 0.88 | 0.75 | 0.247 | 0.329 | 0.46 | 1.22 | 0.74 | 0.220 | 0.293 | 0.45 | 1.20 | 0.51 | 0.002 | 0.006 | 0.33 | 0.77 | 0.51 | 0.002 | 0.008 | 0.34 | 0.78 |
| Adherence to Australian smoking guidelines (non-smokers vs current/former smokers) | 0.80 | 0.365 | 0.438 | 0.50 | 1.29 | 0.80 | 0.355 | 0.397 | 0.78 | 2.01 | 0.82 | 0.503 | 0.503 | 0.45 | 1.48 | 0.83 | 0.572 | 0.572 | 0.43 | 1.60 | 0.79 | 0.449 | 0.490 | 0.42 | 1.46 | 0.76 | 0.364 | 0.397 | 0.43 | 1.36 |

#### Tables 4ab: *Association of lifestyle variables with risk of depression and anxiety over 8 weeks by arm*

| Depression | Full Sample: Unadjusted | | | | | Full Sample: Adjusted for age, sex and SES | | | | | Psychotherapy: Unadjusted | | | | | Psychotherapy: Adjusted for age, sex, and SES | | | | | Lifestyle Therapy: Unadjusted | | | | | Lifetyle Therapy: Adjusted for age, sex, and SES | | | | |
|---|---|---|---|---|---|---|---|---|---|---|---|---|---|---|---|---|---|---|---|---|---|---|---|---|---|---|---|---|---|---|
| | RR | P>\|t\| | Q | L95% CI | U95% CI | RR | P>\|t\| | Q | L95% CI | U95% CI | RR | P>\|t\| | Q | L95% CI | U95% CI | RR | P>\|t\| | Q | L95% CI | U95% CI | RR | P>\|t\| | Q | L95% CI | U95% CI | RR | P>\|t\| | Q | L95% CI | U95%CI |
| Diet* | | | | | | | | | | | | | | | | | | | | | | | | | | | | | | |
| Fruit per 150g increase | 0.90 | 0.080 | 0.216 | 0.79 | 1.01 | 0.90 | 0.085 | 0.287 | 0.79 | 1.01 | 0.91 | 0.233 | 0.373 | 0.78 | 1.06 | 0.91 | 0.256 | 0.458 | 0.77 | 1.07 | 0.89 | 0.208 | 0.373 | 0.74 | 1.07 | 0.89 | 0.255 | 0.458 | 0.74 | 1.08 |
| Vegetable per 75g increase | 0.93 | 0.010 | 0.059 | 0.87 | 0.98 | 0.92 | 0.010 | 0.119 | 0.87 | 0.98 | 0.92 | 0.011 | 0.059 | 0.86 | 0.98 | 0.91 | 0.017 | 0.135 | 0.85 | 0.98 | 0.94 | 0.229 | 0.373 | 0.86 | 1.04 | 0.94 | 0.172 | 0.372 | 0.85 | 1.03 |
| Legume per 150g increase | 0.79 | 0.037 | 0.125 | 0.64 | 0.99 | 0.81 | 0.077 | 0.287 | 0.64 | 1.02 | 0.79 | 0.024 | 0.093 | 0.64 | 0.97 | 0.81 | 0.094 | 0.299 | 0.64 | 1.04 | 0.82 | 0.256 | 0.387 | 0.58 | 1.15 | 0.82 | 0.276 | 0.466 | 0.58 | 1.17 |
| Wholegrain per 80g increase | 0.56 | 0.121 | 0.284 | 0.27 | 1.17 | 0.52 | 0.084 | 0.287 | 0.25 | 1.09 | 0.53 | 0.230 | 0.373 | 0.19 | 1.49 | 0.61 | 0.354 | 0.546 | 0.21 | 1.76 | 0.56 | 0.258 | 0.387 | 0.20 | 1.54 | 0.48 | 0.171 | 0.372 | 0.17 | 1.38 |

| | | | | | | | | | | | | | | | | | | | | | | | | | | | | | | |
|---|---|---|---|---|---|---|---|---|---|---|---|---|---|---|---|---|---|---|---|---|---|---|---|---|---|---|---|---|---|---|
| Nuts and seeds per 30g increase | 0.78 | 0.015 | 0.066 | 0.64 | 0.95 | 0.80 | 0.038 | 0.171 | 0.65 | 0.99 | 0.77 | 0.034 | 0.122 | 0.61 | 0.98 | 0.80 | 0.082 | 0.287 | 0.63 | 1.03 | 0.83 | 0.230 | 0.373 | 0.61 | 1.13 | 0.83 | 0.263 | 0.458 | 0.60 | 1.15 |
| Milk per 250g increase | 0.92 | 0.479 | 0.616 | 0.74 | 1.15 | 0.96 | 0.710 | 0.852 | 0.77 | 1.19 | 0.92 | 0.543 | 0.662 | 0.70 | 1.21 | 0.91 | 0.460 | 0.651 | 0.70 | 1.17 | 0.91 | 0.568 | 0.667 | 0.66 | 1.25 | 0.99 | 0.943 | 0.979 | 0.69 | 1.41 |
| Red meat per 65g increase | 0.98 | 0.881 | 0.898 | 0.80 | 1.21 | 1.01 | 0.894 | 0.947 | 0.82 | 1.25 | 1.03 | 0.844 | 0.876 | 0.78 | 1.35 | 1.02 | 0.869 | 0.939 | 0.78 | 1.35 | 0.93 | 0.618 | 0.695 | 0.71 | 1.23 | 0.97 | 0.829 | 0.915 | 0.74 | 1.28 |
| Processed meat per 50g increase | 1.48 | 0.009 | 0.059 | 1.10 | 1.97 | 1.38 | 0.038 | 0.171 | 1.02 | 1.87 | 1.45 | 0.044 | 0.140 | 1.01 | 2.09 | 1.32 | 0.172 | 0.372 | 0.89 | 1.95 | 1.44 | 0.054 | 0.162 | 0.99 | 2.09 | 1.38 | 0.107 | 0.321 | 0.93 | 2.06 |
| Sugar-sweetened beverage per 375g increase | 1.13 | 0.212 | 0.373 | 0.93 | 1.37 | 1.08 | 0.470 | 0.651 | 0.88 | 1.33 | 1.18 | 0.110 | 0.270 | 0.96 | 1.44 | 1.13 | 0.302 | 0.480 | 0.90 | 1.43 | 1.04 | 0.802 | 0.849 | 0.78 | 1.39 | 1.00 | 0.988 | 0.988 | 0.73 | 1.37 |
| Ultra-processed food per 90g increase | 1.03 | 0.002 | 0.054 | 1.01 | 1.06 | 1.03 | 0.010 | 0.119 | 1.01 | 1.05 | 1.03 | 0.006 | 0.059 | 1.01 | 1.06 | 1.03 | 0.026 | 0.156 | 1.00 | 1.06 | 1.03 | 0.067 | 0.190 | 1.00 | 1.06 | 1.03 | 0.114 | 0.324 | 0.99 | 1.06 |
| Fibre per 30g increase | 0.68 | 0.011 | 0.059 | 0.50 | 0.91 | 0.68 | 0.018 | 0.135 | 0.50 | 0.94 | 0.67 | 0.011 | 0.059 | 0.49 | 0.91 | 0.67 | 0.030 | 0.162 | 0.47 | 0.96 | 0.73 | 0.183 | 0.366 | 0.46 | 1.16 | 0.73 | 0.190 | 0.384 | 0.45 | 1.17 |
| Calcium per 1g increase | 0.74 | 0.235 | 0.373 | 0.46 | 1.21 | 0.81 | 0.443 | 0.651 | 0.48 | 1.38 | 0.80 | 0.454 | 0.598 | 0.44 | 1.45 | 0.87 | 0.643 | 0.806 | 0.47 | 1.59 | 0.71 | 0.366 | 0.507 | 0.34 | 1.48 | 0.76 | 0.502 | 0.678 | 0.34 | 1.71 |
| Polyunsaturated fat (total) per 10% increase in energy | 0.73 | 0.450 | 0.598 | 0.33 | 1.65 | 0.89 | 0.777 | 0.894 | 0.39 | 2.01 | 0.73 | 0.552 | 0.662 | 0.26 | 2.05 | 0.99 | 0.984 | 0.988 | 0.36 | 2.70 | 0.78 | 0.684 | 0.739 | 0.24 | 2.58 | 0.84 | 0.778 | 0.894 | 0.24 | 2.90 |
| Omega-3 per 1g increase | 0.61 | 0.128 | 0.288 | 0.32 | 1.15 | 0.66 | 0.192 | 0.384 | 0.35 | 1.24 | 0.73 | 0.509 | 0.639 | 0.29 | 1.84 | 0.82 | 0.657 | 0.806 | 0.34 | 1.99 | 0.53 | 0.154 | 0.333 | 0.22 | 1.27 | 0.51 | 0.167 | 0.372 | 0.19 | 1.33 |
| Omega-6 per 2% increase in energy | 1.01 | 0.658 | 0.725 | 0.97 | 1.05 | 1.01 | 0.526 | 0.693 | 0.97 | 1.05 | 1.01 | 0.595 | 0.684 | 0.97 | 1.06 | 1.01 | 0.628 | 0.806 | 0.96 | 1.06 | 1.00 | 0.976 | 0.976 | 0.95 | 1.05 | 1.01 | 0.830 | 0.915 | 0.95 | 1.06 |
| Sodium per 2g increase | 1.65 | 0.086 | 0.221 | 0.93 | 2.94 | 1.48 | 0.165 | 0.372 | 0.85 | 2.56 | 1.50 | 0.343 | 0.487 | 0.65 | 3.46 | 1.35 | 0.453 | 0.651 | 0.61 | 2.99 | 1.75 | 0.176 | 0.366 | 0.78 | 3.93 | 1.55 | 0.295 | 0.480 | 0.68 | 3.56 |
| Alcohol | | | | | | | | | | | | | | | | | | | | | | | | | | | | | | |
| Per 1 standard drink per week increase | 1.01 | 0.000 | 0.000 | 1.01 | 1.02 | 1.01 | 0.001 | 0.054 | 1.00 | 1.02 | 1.01 | 0.007 | 0.059 | 1.00 | 1.02 | 1.01 | 0.138 | 0.372 | 1.00 | 1.02 | 1.01 | 0.003 | 0.054 | 1.00 | 1.02 | 1.01 | 0.004 | 0.108 | 1.00 | 1.02 |
| Physical Activity | | | | | | | | | | | | | | | | | | | | | | | | | | | | | | |
| Per 1 hour per week increase | 0.93 | 0.016 | 0.066 | 0.88 | 0.99 | 0.93 | 0.011 | 0.119 | 0.88 | 0.98 | 0.96 | 0.308 | 0.450 | 0.89 | 1.04 | 0.96 | 0.219 | 0.422 | 0.89 | 1.03 | 0.90 | 0.014 | 0.066 | 0.83 | 0.98 | 0.91 | 0.020 | 0.135 | 0.83 | 0.98 |

| Anxiety | Full Sample: Unadjusted | | | | | Full Sample: Adjusted for age, sex and SES | | | | | Psychotherapy: Unadjusted | | | | | Psychotherapy: Adjusted for age, sex, and SES | | | | | Lifestyle Therapy: Unadjusted | | | | | Lifetyle Therapy: Adjusted for age, sex, and SES | | | | |
|---|---|---|---|---|---|---|---|---|---|---|---|---|---|---|---|---|---|---|---|---|---|---|---|---|---|---|---|---|---|---|
| | RR | P>\|t\| | Q | L95% CI | U95% CI | RR | P>\|t\| | Q | L95% CI | U95% CI | RR | P>\|t\| | Q | L95% CI | U95% CI | RR | P>\|t\| | Q | L95% CI | U95% CI | RR | P>\|t\| | Q | L95% CI | U95% CI | RR | P>\|t\| | Q | L95% CI | U95%CI |
| Diet* | | | | | | | | | | | | | | | | | | | | | | | | | | | | | | |
| Fruit per 150g increase | 0.86 | 0.008 | 0.029 | 0.77 | 0.96 | 0.86 | 0.012 | 0.046 | 0.76 | 0.97 | 0.88 | 0.068 | 0.130 | 0.77 | 1.01 | 0.87 | 0.078 | 0.162 | 0.74 | 1.02 | 0.84 | 0.065 | 0.130 | 0.69 | 1.01 | 0.84 | 0.083 | 0.166 | 0.70 | 1.02 |
| Vegetable per 75g increase | 0.90 | 0.000 | 0.000 | 0.86 | 0.96 | 0.90 | 0.001 | 0.008 | 0.84 | 0.95 | 0.90 | 0.001 | 0.007 | 0.84 | 0.95 | 0.88 | 0.001 | 0.008 | 0.82 | 0.95 | 0.91 | 0.060 | 0.125 | 0.83 | 1.00 | 0.91 | 0.046 | 0.113 | 0.83 | 1.00 |
| Legume per 150g increase | 0.73 | 0.006 | 0.023 | 0.59 | 0.92 | 0.73 | 0.014 | 0.050 | 0.57 | 0.94 | 0.73 | 0.005 | 0.021 | 0.59 | 0.91 | 0.71 | 0.018 | 0.061 | 0.53 | 0.94 | 0.74 | 0.070 | 0.130 | 0.53 | 1.02 | 0.75 | 0.093 | 0.179 | 0.53 | 1.05 |
| Wholegrain per 80g increase | 0.60 | 0.265 | 0.388 | 0.24 | 1.48 | 0.55 | 0.211 | 0.326 | 0.22 | 1.41 | 0.51 | 0.289 | 0.411 | 0.15 | 1.77 | 0.49 | 0.262 | 0.393 | 0.14 | 1.71 | 0.69 | 0.540 | 0.663 | 0.21 | 2.25 | 0.59 | 0.373 | 0.480 | 0.19 | 1.88 |
| Nuts and seeds per 30g increase | 0.71 | 0.002 | 0.009 | 0.58 | 0.88 | 0.71 | 0.003 | 0.018 | 0.57 | 0.89 | 0.69 | 0.013 | 0.044 | 0.51 | 0.93 | 0.67 | 0.012 | 0.046 | 0.49 | 0.91 | 0.74 | 0.050 | 0.113 | 0.55 | 1.00 | 0.74 | 0.059 | 0.127 | 0.54 | 1.01 |
| Milk per 250g increase | 0.93 | 0.604 | 0.679 | 0.71 | 1.22 | 0.98 | 0.900 | 0.935 | 0.75 | 1.29 | 0.92 | 0.604 | 0.679 | 0.66 | 1.27 | 0.95 | 0.781 | 0.855 | 0.69 | 1.33 | 0.94 | 0.748 | 0.808 | 0.65 | 1.37 | 1.01 | 0.953 | 0.971 | 0.69 | 1.48 |
| Red meat per 65g increase | 0.54 | 0.000 | 0.000 | 0.39 | 0.76 | 0.55 | 0.000 | 0.000 | 0.39 | 0.76 | 0.66 | 0.101 | 0.176 | 0.40 | 1.08 | 0.67 | 0.107 | 0.193 | 0.40 | 1.09 | 0.43 | 0.000 | 0.000 | 0.27 | 0.69 | 0.43 | 0.000 | 0.000 | 0.27 | 0.69 |
| Processed meat per 50g increase | 1.09 | 0.448 | 0.576 | 0.88 | 1.35 | 1.11 | 0.349 | 0.460 | 0.89 | 1.40 | 1.22 | 0.134 | 0.226 | 0.94 | 1.57 | 1.24 | 0.129 | 0.225 | 0.94 | 1.65 | 0.99 | 0.967 | 0.967 | 0.73 | 1.36 | 1.03 | 0.826 | 0.875 | 0.76 | 1.40 |
| Sugar-sweetened beverage per 375g increase | 1.65 | 0.000 | 0.000 | 1.26 | 2.17 | 1.66 | 0.001 | 0.008 | 1.24 | 2.21 | 1.59 | 0.019 | 0.054 | 1.08 | 2.35 | 1.68 | 0.021 | 0.067 | 1.08 | 2.61 | 1.71 | 0.001 | 0.007 | 1.24 | 2.36 | 1.68 | 0.001 | 0.008 | 1.23 | 2.31 |
| Ultra-processed food per 90g increase | 1.26 | 0.021 | 0.057 | 1.04 | 1.54 | 1.23 | 0.048 | 0.113 | 1.00 | 1.52 | 1.29 | 0.030 | 0.077 | 1.03 | 1.61 | 1.29 | 0.035 | 0.095 | 1.02 | 1.63 | 1.23 | 0.145 | 0.230 | 0.93 | 1.64 | 1.19 | 0.271 | 0.396 | 0.87 | 1.63 |

| | | | | | | | | | | | | | | | | | | | | | | | | | | | | | | |
|---|---|---|---|---|---|---|---|---|---|---|---|---|---|---|---|---|---|---|---|---|---|---|---|---|---|---|---|---|---|---|
| Fibre per 30g increase | 1.04 | 0.000 | 0.000 | 1.02 | 1.06 | 1.04 | 0.001 | 0.008 | 1.01 | 1.06 | 1.04 | 0.014 | 0.044 | 1.01 | 1.06 | 1.04 | 0.023 | 0.069 | 1.00 | 1.07 | 1.04 | 0.002 | 0.009 | 1.02 | 1.07 | 1.04 | 0.007 | 0.032 | 1.01 | 1.07 |
| Calcium per 1g increase | 0.61 | 0.001 | 0.007 | 0.46 | 0.82 | 0.61 | 0.002 | 0.014 | 0.44 | 0.84 | 0.61 | 0.002 | 0.009 | 0.45 | 0.84 | 0.57 | 0.007 | 0.032 | 0.38 | 0.86 | 0.63 | 0.037 | 0.087 | 0.40 | 0.97 | 0.64 | 0.047 | 0.113 | 0.41 | 0.99 |
| Polyunsaturated fat (total) per 10% increase in energy | 0.63 | 0.059 | 0.125 | 0.39 | 1.02 | 0.66 | 0.099 | 0.184 | 0.40 | 1.08 | 0.72 | 0.318 | 0.440 | 0.38 | 1.36 | 0.73 | 0.346 | 0.460 | 0.37 | 1.41 | 0.52 | 0.093 | 0.167 | 0.24 | 1.12 | 0.55 | 0.154 | 0.260 | 0.24 | 1.25 |
| Omega-3 per 1g increase | 0.72 | 0.500 | 0.628 | 0.27 | 1.89 | 0.75 | 0.560 | 0.687 | 0.28 | 2.00 | 0.73 | 0.634 | 0.699 | 0.20 | 2.66 | 0.76 | 0.679 | 0.792 | 0.20 | 2.87 | 0.65 | 0.569 | 0.679 | 0.15 | 2.88 | 0.66 | 0.591 | 0.709 | 0.15 | 3.00 |
| Omega-6 per 2% increase in energy | 0.28 | 0.002 | 0.009 | 0.12 | 0.64 | 0.29 | 0.006 | 0.032 | 0.12 | 0.70 | 0.29 | 0.036 | 0.087 | 0.09 | 0.92 | 0.29 | 0.050 | 0.113 | 0.09 | 1.00 | 0.27 | 0.017 | 0.051 | 0.09 | 0.79 | 0.29 | 0.030 | 0.085 | 0.09 | 0.89 |
| Sodium per 2g increase | 1.02 | 0.416 | 0.548 | 0.98 | 1.06 | 1.02 | 0.286 | 0.396 | 0.98 | 1.06 | 1.03 | 0.266 | 0.388 | 0.98 | 1.07 | 1.03 | 0.205 | 0.326 | 0.98 | 1.08 | 1.01 | 0.869 | 0.902 | 0.95 | 1.07 | 1.01 | 0.689 | 0.792 | 0.96 | 1.07 |
| Alcohol | | | | | | | | | | | | | | | | | | | | | | | | | | | | | | |
| Per 1 standard drink per week increase | 1.00 | 0.588 | 0.679 | 0.99 | 1.01 | 1.00 | 0.707 | 0.795 | 0.99 | 1.01 | 1.01 | 0.371 | 0.501 | 0.99 | 1.02 | 1.01 | 0.525 | 0.659 | 0.99 | 1.02 | 1.00 | 0.948 | 0.966 | 0.99 | 1.01 | 1.00 | 0.991 | 0.991 | 0.99 | 1.01 |
| Physical Activity | | | | | | | | | | | | | | | | | | | | | | | | | | | | | | |
| Per 1 hour per week increase | 0.96 | 0.262 | 0.388 | 0.90 | 1.03 | 0.97 | 0.285 | 0.396 | 0.91 | 1.03 | 0.99 | 0.818 | 0.866 | 0.91 | 1.08 | 0.99 | 0.792 | 0.855 | 0.91 | 1.08 | 0.93 | 0.140 | 0.229 | 0.85 | 1.02 | 0.94 | 0.178 | 0.291 | 0.85 | 1.03 |

*Dietary components recorded in grams were additionally adjusted for energy intake using Willett's residual method
